# A genome wide association study to identify germline variants associated with cancer-associated cachexia - a preliminary analysis

**DOI:** 10.1101/2023.07.24.23293083

**Authors:** Ashok Narasimhan, Mahalakshmi Kumaran, Ioannis Gioulbasanis, Richard J E Skipworth, Oliver F Bathe, Stein Kaasa, Florian Strasser, Bruno Gagnon, Vickie Baracos, Sambasivarao Damaraju

## Abstract

**Background:** Cancer cachexia is characterized by severe loss of muscle and fat involving a complex interplay of host-tumor interactions. While much emphasis has been made in understanding the molecular mechanisms associated with cachexia, understanding the heritable component of cachexia remains less explored. The current study aims to identify Copy Number Variants (CNV) as genetic susceptibility determinants for weight loss in patients with cancer cachexia using genome wide association study (GWAS) approach.

**Methods:** A total of 174 age-matched patients with esophagogastric or lung cancer were classified as weight losing (>10% weight loss) or weight stable participants (<2% weight loss). DNA was genotyped using Affymetrix SNP 6.0 arrays to profile CNVs. We tested CNVs with >5% frequency in the population for association with weight loss. Pathway analysis was performed using the genes embedded within CNVs. To understand if the CNVs in the present study are also expressed in skeletal muscle of patients with cachexia, we utilized two publicly available human gene expression datasets to infer the relevance of identified genes in the context of cachexia.

**Results:** Among the associated CNVs, 5414 CNVs had embedded protein coding genes. Of these, 1583 CNVs were present at >5% frequency. We combined multiple contiguous CNVs within the same genomic region and called them Copy Number Variable Region (CNVR). This led to identifying 896 non-redundant CNV/CNVRs which encompassed 803 protein coding genes. Genes embedded within CNVs were enriched for several pathways implicated in cachexia and muscle wasting including JAK-STAT signaling, Oncostatin M signaling, Wnt signaling and PI3K-Akt signaling. This is the first proof of principle GWAS study to identify CNVs as genetic determinants for cancer cachexia. Further, we show that a subset of CNV/CNVR embedded genes identified in the current study are common with the previously published skeletal muscle gene expression datasets, indicating that expression of CNV/CNVR genes in muscle may have functional consequences in patients with cachexia These genes include CPT1B, SPON1, LOXL1, NFAT5, RBFOX1 and PCSK6 to name a few.

**Conclusion:** This is the first proof of principle GWAS study to identify CNVs as genetic determinants for cancer cachexia. The data generated will aid in future replication studies in larger cohorts to account for genetic susceptibility to weight loss in patients with cancer cachexia.

## Introduction

Cancer-associated cachexia, which is characterized by involuntary weight loss, is evident in majority of patients with cancer in their advanced stages of the disease and is responsible for approximately 30% of cancer-related deaths ^1–3^. Cancer cachexia shows considerable variation in both prevalence and severity^4,5^. Some patients are unaffected whereas others become profoundly wasted. Based on our current knowledge, we are unable to predict, for any given cohort of patients, who will develop cancer cachexia and who will not. Certain tumor primary sites (e.g., pancreatic, gastrointestinal and lung) and advanced tumor stage at diagnosis are factors associated with higher cachexia prevalence^2^. While cachexia is in part directly driven by tumor, patient-level susceptibility to wasting may also contribute to inter-individual variability.

Identifying and accounting for heritability in several diseases/traits using germline DNA has been the subject of intense investigations^6^. Single Nucleotide Polymorphisms (SNPs) are often studied as genomic markers^7,8^ in genome-wide association studies (GWAS). Germline SNPs are now well-established genetic determinants of body weight, muscle mass and strength, fat mass, insulin resistance, inflammation and, therefore, in all likelihood a determinant of cachexia^9^. Previous candidate SNP studies showed associations with weight loss in patients with cancer cachexia^10,11^. However, SNP markers are predominantly in the intergenic regions, are not readily amenable for biological interpretations and require fine mapping of associated regions to identify the causal genes or regulatory domains. On the other hand, Copy Number Variants (CNVs) are structural variants that encompass large genomic regions with embedded coding and non-coding gene regions. CNVs are also currently targeted as markers of choice in GWAS^12^. CNVs include amplifications or deletions of DNA and may also confer gene dosage effects. CNVs range from 50 base pairs (bp) to more than 1 mega base pairs (Mbs) in length^13^. Cumulative proportion of genome covered by CNVs is greater than SNPs, which may have a profound impact on the phenotype^14,15^. CNVs can also regulate expression of genes through cis or trans effects ^16,17^. We focus on profiling CNVs as potential genetic determinants for cancer associated cachexia using a GWAS approach to facilitate identification of individuals who are at risk for developing cachexia early in their disease trajectory and for possible stratification of patients for therapeutic interventions. We hypothesize that weight loss severity in patients with cancer is heritable and the heritability is explained in part through the association of common CNV polymorphisms. Specific objectives in the present study are to:

i. Identify germline common CNVs from patients with cancer, with and without weight loss as distinct traits.
ii. Gain biological insights by analyzing the protein coding genes embedded within CNVs through in-silico pathway analytic approaches.
iii. Investigate if the genes embedded within CNVs are also present as differentially expressed genes in skeletal muscle of patients with cancer cachexia.

## Materials and Methods

### Procurement of samples and Isolation of DNA

In this preliminary study, we utilized a subset of samples from our previously published candidate SNP study in cachexia^10,11^ in which weight loss history and germline DNA were sampled (n=1276) from patients at risk for cachexia with United Kingdom, Canada, Norway, Switzerland, and Greece as participating countries. We limited our investigation to two primary cancers (lung and gastro-esophageal) for their relative higher prevalence of cachexia in patients^2^. We classified individuals based on the cachexia consensus definition of weight loss to stratify patients as cases and controls. This being a preliminary study, we selected individuals with extremes of weight loss of >10% over a period of six months (hereafter termed cases) for comparison with weight stable patients (<2% weight loss in six months, hereafter termed controls). A total of 179 patients met these criteria (74 cachectic cases and 105 controls) and were included in the study for genotyping. Complete demographics, clinical information, inclusion, and exclusion criteria of the study participants from multiple cancer types are provided elsewhere^10,11^. All patients provided written and informed consent for the study. DNA was extracted from buffy coat and was stored at -80°C until further use. The Health Research Ethics Board of Alberta (HREBA)-Cancer Committee approved the study protocol (# 17-0517).

### Genotyping of samples using SNP 6.0 arrays and quality control

Affymetrix SNP 6.0 arrays contains probes for both SNPs and CNVs totaling 1.8 million genetic markers. As a part of quality control (QC), contrast QC was calculated for each sample and genotyping quality was considered good if the contrast QC was above 1.7, as described in our previous publications^12,18,19^. SNP genotyping calls from the array data were used to identify the genetic ancestry of the cohort we used. Principal component analysis (PCA) was performed using EIGENSTRAT to identify population structure using HapMap 270 genotype data as reference (European, Han Chinese or African as sub populations). The study participants were mapped to identify their genetic ancestry and samples that were not mapping to European ancestry were removed as outliers. Identity by descent (IBD) analysis was also performed to identify cryptic relatedness [16].

## Identification of CNVs

CNV analysis was performed using Partek Genomics Suite v6.6 [PGS, (Partek® Genomics Suite software, Version 6.6 beta, Copyright © 2009 Partek Inc., St. Louis, MO, USA)]. CNV intensity files from hybridized arrays were imported and the default parameters such as GC wave corrections were used. We used all sample normalization approach to create a reference baseline and to calculate the copy number estimates for each sample. Briefly, average hybridization intensities from all samples and all probes were treated as a representative of a diploid genome as described previously^12,18,20^. To identify the copy number status and define the CNV boundaries, genomic segmentation was carried out using the default parameters in PGS: minimum consecutive genomic markers > 10, p-value threshold= 0.001, signal to noise ratio= 0.3. A segment of CNV was called copy gain when the intensity value was above 2.3 and called copy loss if the intensity value was below 1.7, a range optimized for genomic segmentation algorithm in a diploid region. A copy number was assigned a diploid status when the intensity values fell in the range of 1.7 and 2.3. CNVs with p < 0.05 was considered significant (this p-value is distinct from the p-value in the genomic segmentation). The PGS output for CNV is given as contiguous segments with frequency and p-value calculated for each CNV call. The nature of the segmentation algorithm is that the CNV calls are made for stretches of DNA and the number of probes present along the linear length of the chromosome. As a result, some of the CNVs vary in length; multiple CNVs within the same genomic region were joined to form a contiguous segment and were referred to as a single Copy Number Variable Region (CNVR) as described in our previous studies^18^. As CNVs harbor both coding and non-coding regions of the genome, we restricted our present analysis to map CNVs or CNVRs that showed 100 % overlap with the protein coding genes for a detailed analysis. The non-coding genes were also catalogued, as these may confer regulatory roles and await further studies to identify their significance. Genome build hg19 was used as the reference genome for mapping CNVs. CNVs with a total frequency of 5% or more were defined as common CNVs^12,18,20^.

In addition, we also mapped the significant CNV/CNVRs to 1000 Genomes Project phase 3 data and Database of Genomic Variants (DGV, http://dgv.tcag.ca/dgv/app/home) to interrogate if the identified CNV/CNVRs are recorded as common variants at a population level (distinct from de novo variants) in well annotated independent structural variation databases. Data deposited into such databases utilize independent CNV calling algorithms and independent genotyping platforms. Overlap of variants identified in this study to those from the curated databases may be viewed as an independent validation of CNVs identified in the current study. Those CNVs/CNVRs not mapping to these databases were considered as potentially novel variants that may require independent validation.

### Biological interpretation of CNV embedded protein coding genes

CNVs which had a 100% overlap with protein coding genes at pre-defined cut-offs were considered for in silico pathway analysis (see results for defining cut-offs). Pathway analysis was performed using Metascape^21^. To identify nodal molecules and their downstream targets, we performed network analysis using STRING database (https://string-db.org/).

To further understand the biological relevance of the identified CNV/CNVRs, we mapped the 803 unique protein coding genes obtained from 896 CNVs and compared them with the publicly available human skeletal muscle gene expression datasets. For this, we utilized differentially expressed genes reported in cancer cachexia studies from two datasets-GSE133979 and GSE18832^22,23^. Both these gene profiling studies utilized non-cancer controls and patients with cancer presented with weight loss. We only considered differentially genes that had a fold change of 1.5 and p <0.05.

## Results

The overall study workflow is presented in Figure 1.

**Figure 1.**
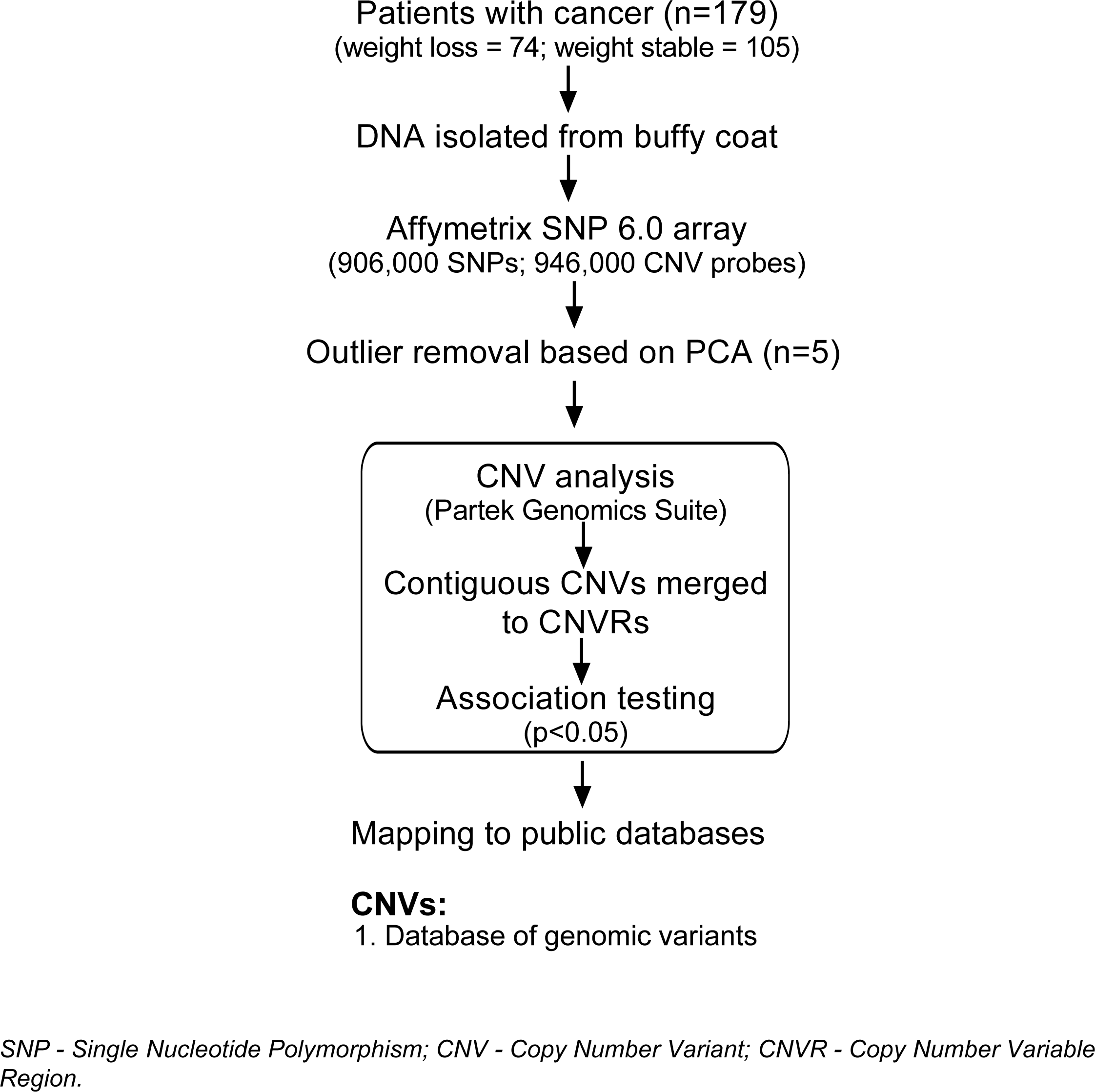
Overall study workflow.

## Demographics of study participants and Quality control of SNP 6.0 arrays

PCA identified 5 outliers (3 cachectic cases and 2 weight stable controls) which were removed from further analysis (see principal component analysis, Figure 2) leaving 71 cases and 103 controls for further analysis. Identity by descent analysis revealed no cryptic relatedness in the samples.

**Figure 2.**
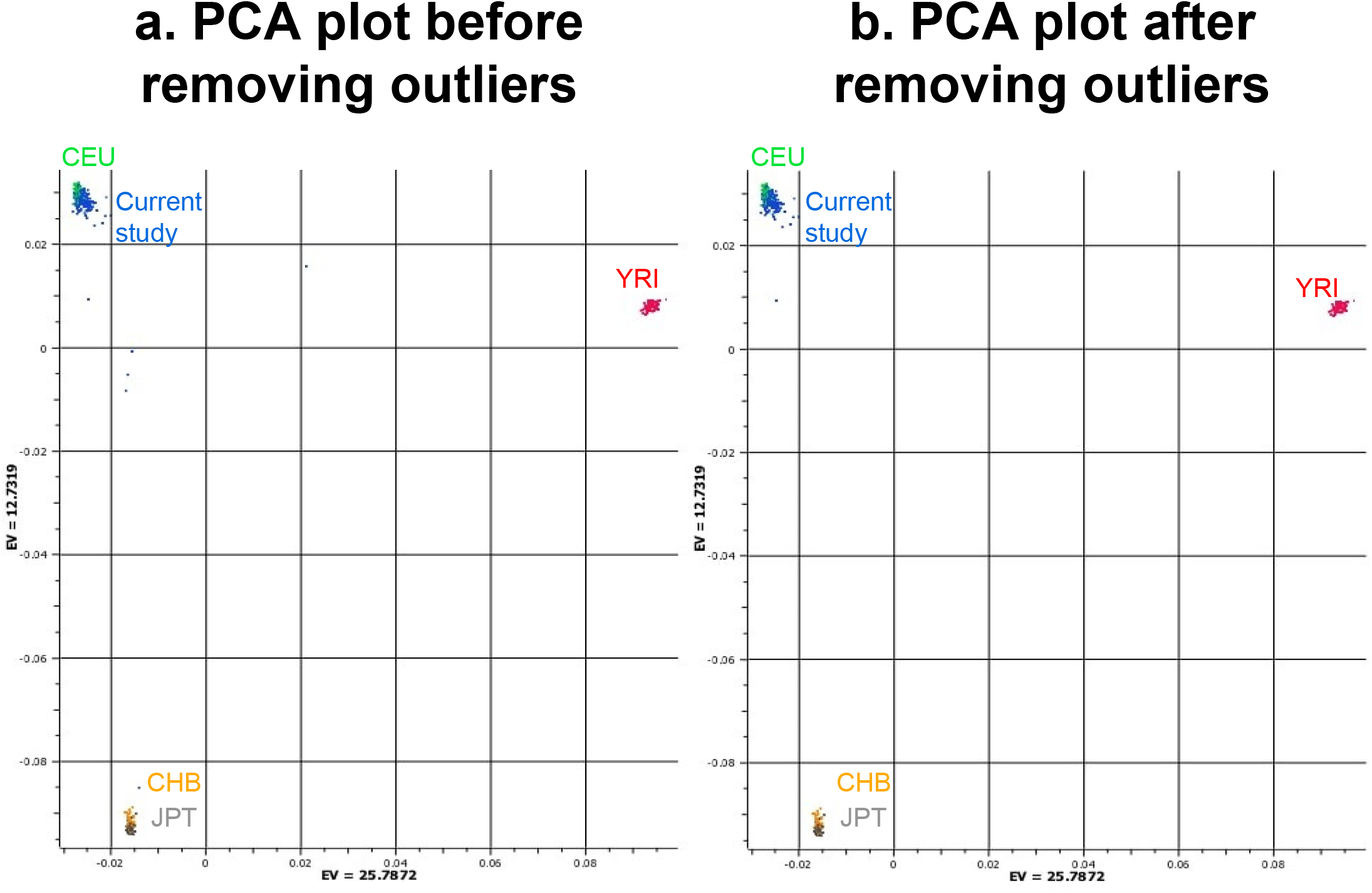
Principal component analysis (PCA) before removing outliers: **A**.The samples from the current study were mapped to HapMap 270 project where Red indicates Yoruba population (YRI), Orange indicates Han Chinese population (CHB), Dark grey (JPT) indicates Japanese population and Green indiates CEU indicates Utah residents with Northern and Western European ancestry from the CEPH collection. Blue indicates samples from the current study which maps with the European ancestry. **B. after removing outliers**: Samples with more than 3 standard deviations were removed from the study as described earlier^19^.

Table 1 summarizes the patient demographics. There was no significant difference for age and sex between weight losing (cases) and weight stable cancer (controls) individuals. The weight losing group showed low BMI compared to weight stable cancer group and the differences were statistically significant. A mean of 15.8% weight loss was observed among cases; control participants did not undergo weight loss. Tumor type was also significantly different between the groups.

**Table 1.**
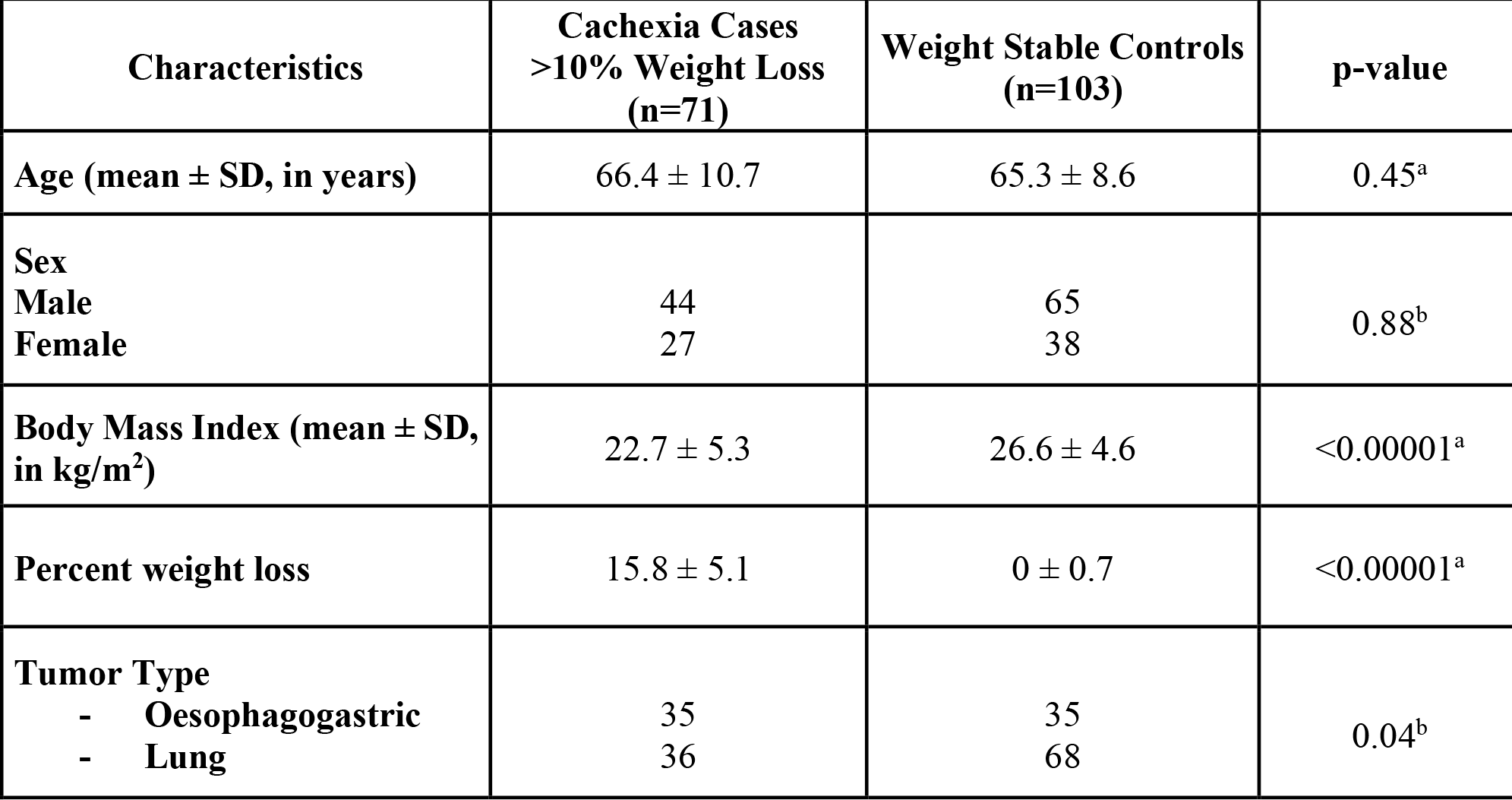
Patient demographics. Values are represented as mean ± standard deviation. ^a^independent t-test, ^b^chi square test. For the current study, we have interchangeably used the terms cancer cachexia and weight loss. The CT image data and quantifications are not available for the cases and controls used in this study; hence sarcopenic status could not be ascertained. SD-Standard deviation

| Characteristics | Cachexia Cases<br>>10% Weight Loss<br>(n=71) | Weight Stable Controls<br>(n=103) | p-value |
| --- | --- | --- | --- |
| Age (mean ± SD, in years) | 66.4 ± 10.7 | 65.3 ± 8.6 | 0.45<sup>a</sup> |
| Sex<br>Male<br>Female | 44<br>27 | 65<br>38 | 0.88<sup>b</sup> |
| Body Mass Index (mean ± SD,<br>in kg/m<sup>2</sup>) | 22.7 ± 5.3 | 26.6 ± 4.6 | <0.00001<sup>a</sup> |
| Percent weight loss | 15.8 ± 5.1 | 0 ± 0.7 | <0.00001<sup>a</sup> |
| Tumor Type<br>- Oesophagogastric<br>- Lung | 35<br>36 | 35<br>68 | 0.04<sup>b</sup> |

### Characterization of CNVs

After performing quality control, and calculating the copy number estimates, genomic segmentation for CNV with predefined cut-off led to the identification of 236,502 CNVs. Since CNVs may encompass both protein coding and non-protein coding genes, we interrogated for all the genes within CNVs (Figure 3A). Approximately, 56% of the genes within CNVs were protein coding genes and the remaining were noncoding RNAs which included miRNAs, lncRNAs, piRNAs, among others. Of these, only miRNAs are well characterized for their gene regulatory functions and the roles of other non-coding RNAs are still evolving^12,18^. The length distribution of CNVs in protein coding region is given in Figure 3B. (See supplementary Table S1 for a complete list of statistically significant CNVs associated across all 22 autosomes). The majority of the CNVs fell between 1-5 kb length. CNVs in X, Y and mitochondrial (MT) chromosomes were filtered resulting in 227,074 CNVs which were subjected for further filtering. At a statistical significance cut-off of at p<0.05, 10,923 CNVs were identified (any bp length). 10,801 CNVs were presented with >50 bp length. From this subset of CNVs, 5414 CNVs were retained when filtered for CNVs that showed 100% overlap with protein coding genes. Of these, 1583 CNVs had more than 5% frequency in the study population.

**Figure 3.**
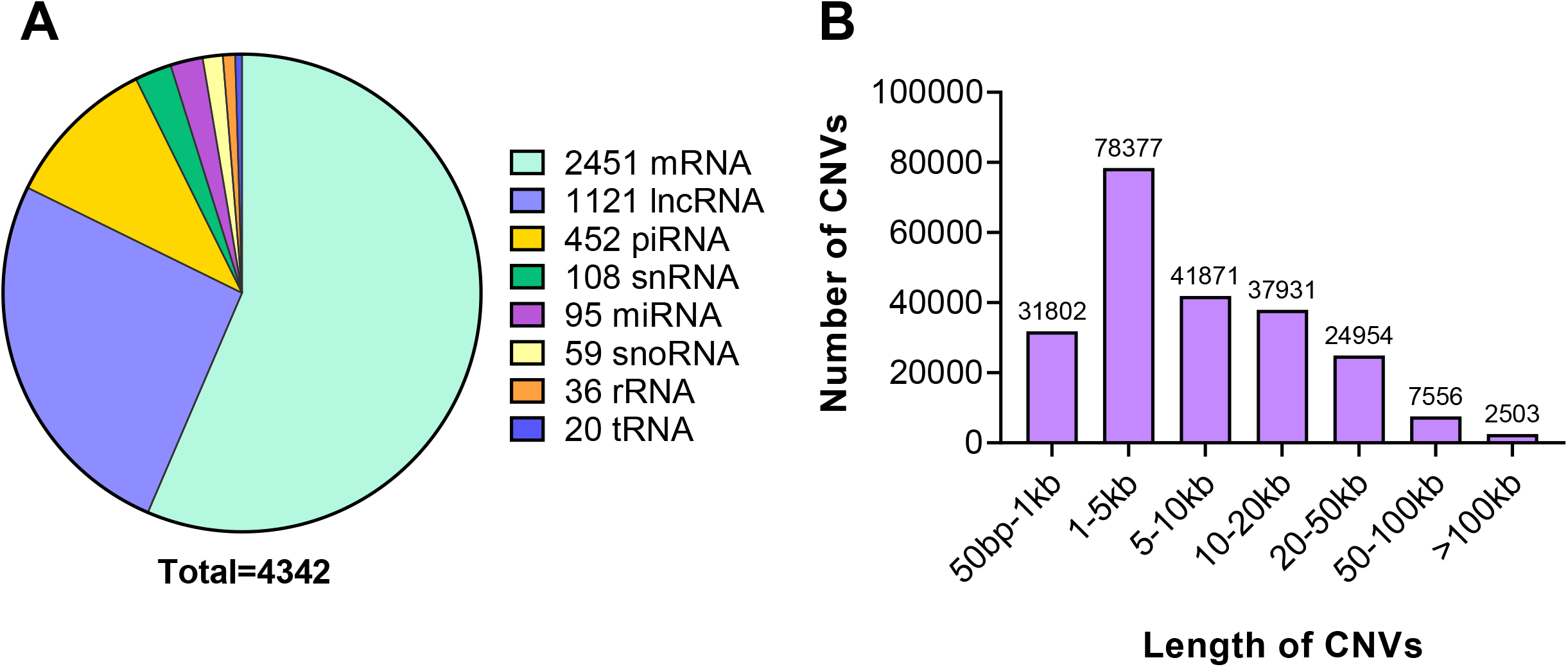
Characterization of CNVs. **A**. CNVs identified in coding and non-coding regions of the genome are shown. CNVs with more than 50 base pairs were retained and CNVs present in X and Y chromosomes were removed. **B**. Distribution of CNVs based on their base pair length where the X-axis represents the length, and the Y axis represents the number of CNVs in each category. mRNA-messenger RNA, lncRNA-long non-coding RNA, piRNA-piwi-interacting RNA, miRNA-microRNA, snoRNA-small nucleolar RNAs, rRNA-ribosomal RNA, tRNA-transfer RNA. CNVs-Copy number variants.

Further, contiguous genomic regions were merged from the 1583 CNVs into copy number variable regions (CNVRs) ^12,18^ CNVs which did not have contiguous regions were not merged and were called as CNVs. In all, 896 non-redundant CNV/CNVRs were identified which embedded 803 unique protein coding genes. Copy gain and loss may occur in the same subject at a given locus in both cases and controls contributing to a redundancy of CNV counts. The cumulative copy number gain regions from 896 CNV/CNVRs, were 4208 and 2492 in cachectic cases and weight stable controls, respectively. The observed copy number loss regions were 4153 and 2774 in cachectic cases and weight stable controls respectively. The observed differences in copy gain/loss in cases and controls were statistically significant (p=0.0006, chi-square test). 743/896 CNV/CNVRs (∼83%) overlapped with 1000 Genomes phase 3 data or DGV and the remaining CNVs may potentially be novel variants requiring further independent validations.

### CNV embedded genes are associated with pathways related to muscle wasting and metabolism

The top 30 CNV/CNVRs with the highest frequency identified in this study are presented in Table 2. To date, several of the identified CNV embedded genes have not been directly implicated in cachexia. A complete list of significant CNVs/CNVRs identified in this study are presented in Supplementary Table S1 and Fig 4A and further data summaries pertaining to pathways and biological networks are represented in Figure 4B. Pathways associated with protein synthesis and atrophy such as PI3K-Akt signaling, Jak-STAT signaling and FOXO signaling were identified (Figure 4B). Representative upstream regulators include RBFOX1, JAK1, JAK2, JAK3 and LIFR (Figure 4C). Many of these genes have been associated with muscle wasting in patients with cancer.

**Table 2.**
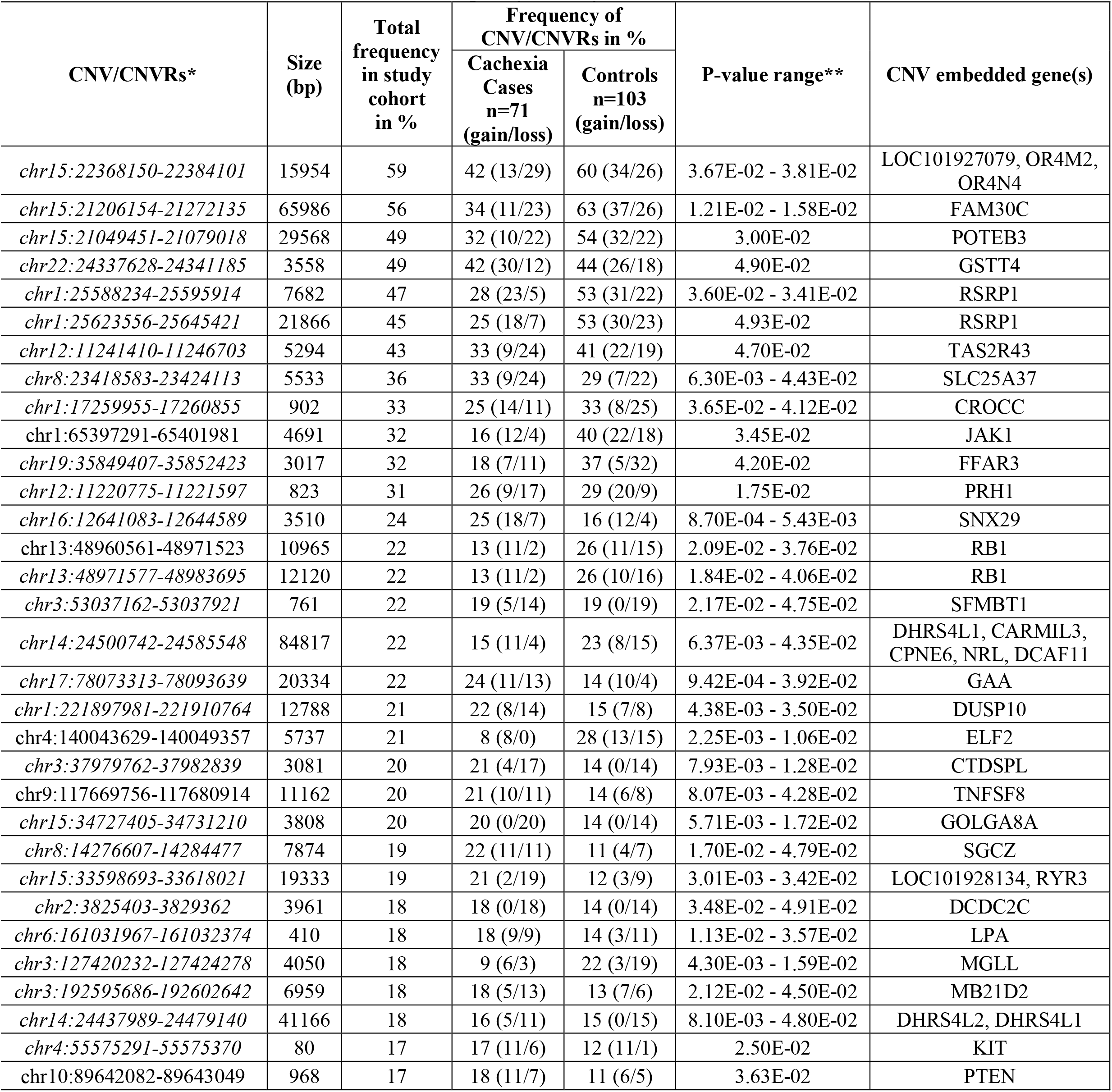
Copy Number Variants/Copy Number Variable Regions associated with cancer cachexia based on frequency in study cohort: *CNV/CNVRs that are italicized indicate that these are also present in the 1000 genomes project or database of genomic variants. **As each CNV has its own p-value, when contiguous CNVs were merged as CNVRs, p-value range for those regions were calculated and shown. CNV-copy number variants, CNVRs-copy number variable regions. The total frequency in % is calculated as a percentage by taking number of aberrations in both copy loss/gain to the total number of samples (n=174). The number of CNV/CNVRs represents the total number of aberrations in case and control.

| CNV/CNVRs* | Size (bp) | Total frequency in study cohort in % | Frequency of CNV/CNVRs in % |  | P-value range** | CNV embedded gene(s) |
| --- | --- | --- | --- | --- | --- | --- |
|  |  |  | Cachexia Cases n=71 (gain/loss) | Controls n=103 (gain/loss) |  |  |
| *chr15:22368150-22384101* | 15954 | 59 | 42 (13/29) | 60 (34/26) | 3.67E-02 - 3.81E-02 | LOC101927079, OR4M2, OR4N4 |
| *chr15:21206154-21272135* | 65986 | 56 | 34 (11/23) | 63 (37/26) | 1.21E-02 - 1.58E-02 | FAM30C |
| *chr15:21049451-21079018* | 29568 | 49 | 32 (10/22) | 54 (32/22) | 3.00E-02 | POTEB3 |
| *chr22:24337628-24341185* | 3558 | 49 | 42 (30/12) | 44 (26/18) | 4.90E-02 | GSTT4 |
| *chr1:25588234-25595914* | 7682 | 47 | 28 (23/5) | 53 (31/22) | 3.60E-02 - 3.41E-02 | RSRP1 |
| *chr1:25623556-25645421* | 21866 | 45 | 25 (18/7) | 53 (30/23) | 4.93E-02 | RSRP1 |
| *chr12:11241410-11246703* | 5294 | 43 | 33 (9/24) | 41 (22/19) | 4.70E-02 | TAS2R43 |
| *chr8:23418583-23424113* | 5533 | 36 | 33 (9/24) | 29 (7/22) | 6.30E-03 - 4.43E-02 | SLC25A37 |
| *chr1:17259955-17260855* | 902 | 33 | 25 (14/11) | 33 (8/25) | 3.65E-02 - 4.12E-02 | CROCC |
| *chr1:65397291-65401981* | 4691 | 32 | 16 (12/4) | 40 (22/18) | 3.45E-02 | JAK1 |
| *chr19:35849407-35852423* | 3017 | 32 | 18 (7/11) | 37 (5/32) | 4.20E-02 | FFAR3 |
| *chr12:11220775-11221597* | 823 | 31 | 26 (9/17) | 29 (20/9) | 1.75E-02 | PRH1 |
| *chr16:12641083-12644589* | 3510 | 24 | 25 (18/7) | 16 (12/4) | 8.70E-04 - 5.43E-03 | SNX29 |
| *chr13:48960561-48971523* | 10965 | 22 | 13 (11/2) | 26 (11/15) | 2.09E-02 - 3.76E-02 | RB1 |
| *chr13:48971577-48983695* | 12120 | 22 | 13 (11/2) | 26 (10/16) | 1.84E-02 - 4.06E-02 | RB1 |
| *chr3:53037162-53037921* | 761 | 22 | 19 (5/14) | 19 (0/19) | 2.17E-02 - 4.75E-02 | SFMBT1 |
| *chr14:24500742-24585548* | 84817 | 22 | 15 (11/4) | 23 (8/15) | 6.37E-03 - 4.35E-02 | DHRS4L1, CARMIL3, CPNE6, NRL, DCAF11 |
| *chr17:78073313-78093639* | 20334 | 22 | 24 (11/13) | 14 (10/4) | 9.42E-04 - 3.92E-02 | GAA |
| *chr1:221897981-221910764* | 12788 | 21 | 22 (8/14) | 15 (7/8) | 4.38E-03 - 3.50E-02 | DUSP10 |
| *chr4:140043629-140049357* | 5737 | 21 | 8 (8/0) | 28 (13/15) | 2.25E-03 - 1.06E-02 | ELF2 |
| *chr3:37979762-37982839* | 3081 | 20 | 21 (4/17) | 14 (0/14) | 7.93E-03 - 1.28E-02 | CTDSPL |
| *chr9:117669756-117680914* | 11162 | 20 | 21 (10/11) | 14 (6/8) | 8.07E-03 - 4.28E-02 | TNFSF8 |
| *chr15:34727405-34731210* | 3808 | 20 | 20 (0/20) | 14 (0/14) | 5.71E-03 - 1.72E-02 | GOLGA8A |
| *chr8:14276607-14284477* | 7874 | 19 | 22 (11/11) | 11 (4/7) | 1.70E-02 - 4.79E-02 | SGCZ |
| *chr15:33598693-33618021* | 19333 | 19 | 21 (2/19) | 12 (3/9) | 3.01E-03 - 3.42E-02 | LOC101928134, RYR3 |
| *chr2:3825403-3829362* | 3961 | 18 | 18 (0/18) | 14 (0/14) | 3.48E-02 - 4.91E-02 | DCDC2C |
| *chr6:161031967-161032374* | 410 | 18 | 18 (9/9) | 14 (3/11) | 1.13E-02 - 3.57E-02 | LPA |
| *chr3:127420232-127424278* | 4050 | 18 | 9 (6/3) | 22 (3/19) | 4.30E-03 - 1.59E-02 | MGLL |
| *chr3:192595686-192602642* | 6959 | 18 | 18 (5/13) | 13 (7/6) | 2.12E-02 - 4.50E-02 | MB21D2 |
| *chr14:24437989-24479140* | 41166 | 18 | 16 (5/11) | 15 (0/15) | 8.10E-03 - 4.80E-02 | DHRS4L2, DHRS4L1 |
| *chr4:55575291-55575370* | 80 | 17 | 17 (11/6) | 12 (11/1) | 2.50E-02 | KIT |
| *chr10:89642082-89643049* | 968 | 17 | 18 (11/7) | 11 (6/5) | 3.63E-02 | PTEN |

**Figure 4.**
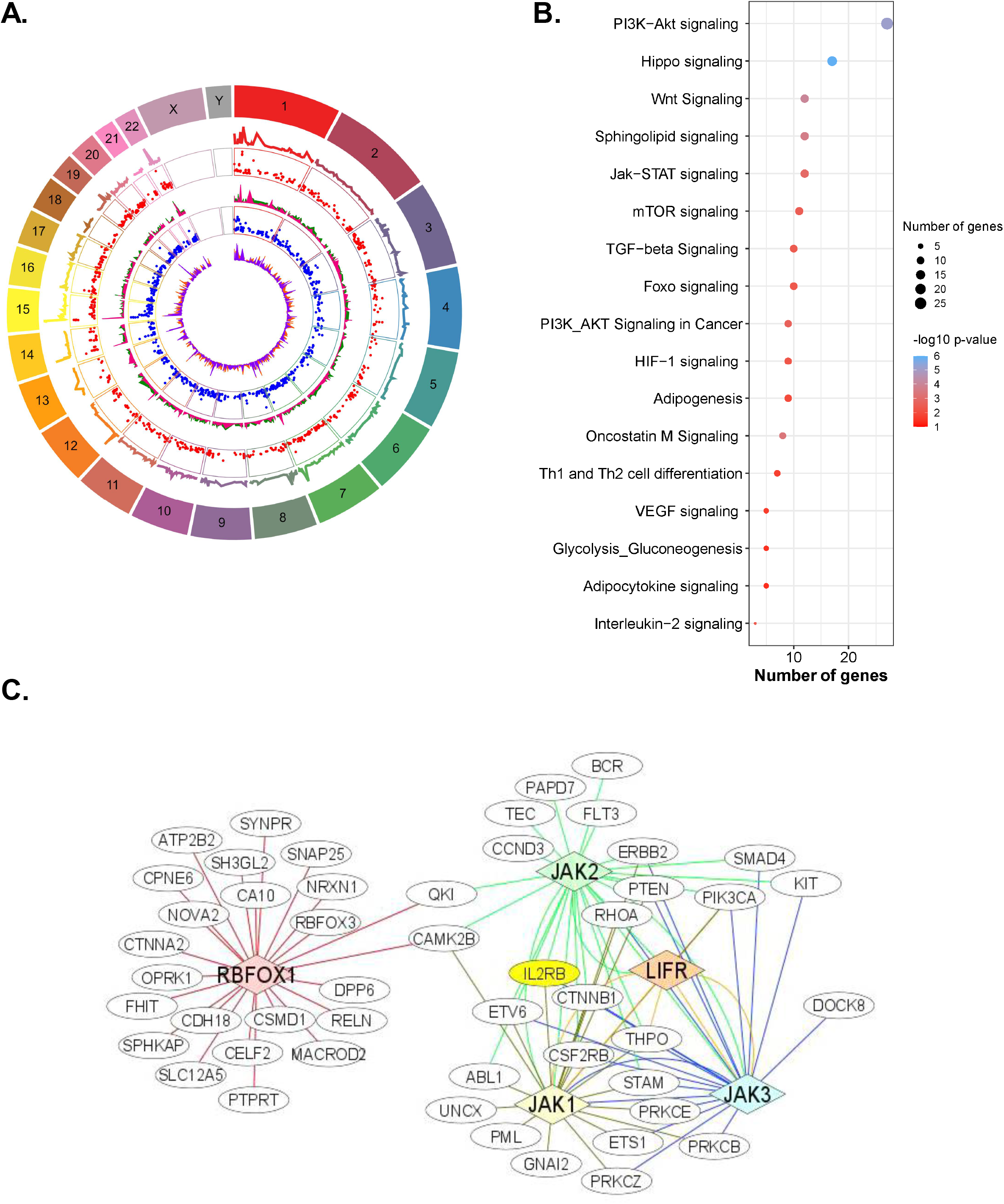
Overlap of CNV/CNVRs embedded protein coding genes. **A**. Circos plot representing the significant 896 CNV/CNVRs with 100% overlap with gene. Track 1 (outermost circle) -chromosome; track 2 – total aberrations represented as line graph. Colors are same as chromosome colors; track 3 – total amplifications represented as scatter plot. Red color indicates amplification; track 4 – has two layers as area graph and represents total number of amplifications in cases (green) and controls (pink); track 5 – total deletions represented as scatter plot. Blue color indicates deletions; track 6 – has two layers as area graph. Represents total number of deletions in cases (orange) and controls (violet). Outlines in tracks 3 and 5 delineate the chromosome boundaries. **B**. Genes embedded in CNVRs were subjected to pathway analysis using Metascape and the pathways are represented in the dot plot. **C**. Genes with more than 5 connections to their neighboring genes are represented. The diamond shape indicates nodal molecules while the circled genes are the downstream genes. CNV-copy number variants, CNVRs-copy number variable regions

### CNV embedded protein coding genes are also differentially expressed in publicly available skeletal muscle datasets

We identified 52 genes which are common between CNV/CNVRs embedded gene dataset (this study) and skeletal muscle gene expression datasets (GSE133979 and GSE18832)^22,23^. Genes such as SPON1, CPT1B, SLC37A2, ARID5B, RBFOX1, ABLIM1 and GALNT15 (Table 3 and Supplementary Table S2) have not been reported to play a role in cachexia pathophysiology in literature. CNVs harbor genes spanning several loci and as such a subset of genes which are also expressed in skeletal muscle gene expression data sets are emphasized. For example, RASSF1 gene which was embedded in chr3:50344972-50454597 was differentially expressed. But CYB561D2 and CACNA2D2 which were present in the same CNV region were not differentially expressed in skeletal muscle. The complete list of CNVs embedded protein coding genes that are differentially expressed in presented in Table S2.

**Table 3.** Genes embedded within CNVs/CNVRs shown to be differentially expressed in skeletal muscle of patients with cancer cachexia. The list of 30 genes represented above are the ones which are differentially expressed in human skeletal muscle tissue transcriptome studies as described in methods. CNV/CNVRs that are italicized (first column in the table) are also present in the 1000 genomes project or database of genomic variants. The total frequency in % is calculated as a percentage by taking number of aberrations in both copy loss/gain to the total number of samples (n=174). The number of CNV/CNVRs represents the total number of aberrations in case and control.

| CNV/CNVRs* | Size (bp) | Total frequency in study cohort in % | Frequency of CNV/CNVRs in % |  | P-value range** | CNV embedded gene(s) |
| --- | --- | --- | --- | --- | --- | --- |
|  |  |  | Cachexia Cases n=71 (gain/loss) | Controls n=103 (gain/loss) |  |  |
| *chr7:143914612-143929685* | 15076 | 41 | 34 (10/24) | 38 (21/17) | 2.03E-02-3.96E-02 | OR2A1-AS1, OR2A42 |
| *chr22:51006226-51016725* | 10502 | 15 | 19 (7/12) | 7 (4/3) | 9.66E-04-1.83E-02 | CPT1B |
| *chr16:16282313-16357764* | 75459 | 14 | 9 (6/3) | 16 (3/13) | 1.43E-02- 4.54E-02 | ABCC6, NOMO3 |
| *chr3:50344972-50454597* | 109636 | 14 | 17 (9/8) | 8 (8/0) | 6.90E-03-4.70E-02 | RASSF1, CYB561D2, CACNA2D2 |
| *chr7:150489319-150501847* | 12530 | 13 | 13 (4/9) | 9 (0/9) | 2.84E-03-3.22E-02 | TMEM176B, TMEM176A |
| *chr21:40190571-40190670* | 100 | 12 | 7 (7/0) | 14 (6/8) | 3.88E-02 | ETS2 |
| *chr3:133009848-133011437* | 1590 | 12 | 13 (0/13) | 8 (1/7) | 4.81E-02 | TMEM108 |
| *chr3:133012075-133014938* | 2865 | 12 | 13 (0/13) | 8 (1/7) | 1.84E-02-4.81E-02 | TMEM108 |
| *chr22:21728129-21805650* | 77524 | 12 | 15 (10/5) | 6 (4/2) | 1.56E-02-1.82E-02 | RIMBP3B, HIC2 |
| *chr19:50353707-50402542* | 48841 | 11 | 14 (7/7) | 6 (5/1) | 7.97E-03-4.35E-02 | PNKP, AKT1S1, TBC1D17, IL4I1 |
| *chr21:40190745-40190820* | 76 | 11 | 7 (7/0) | 12 (5/7) | 4.14E-02 | ETS2 |
| *chr20:62275230-62280003* | 4774 | 10 | 12 (7/5) | 6 (5/1) | 3.70E-02 | STMN3 |
| *chr3:64752341-64754300* | 1960 | 10 | 11 (4/7) | 6 (0/6) | 2.80E-02 | ADAMTS9-AS2 |
| *chr7:100769756-100789977* | 20225 | 9 | 5 (4/1) | 11 (0/11) | 3.88E-03-1.06E-02 | SERPINE1 |
| *chr11:124946298-124947103* | 806 | 9 | 11 (5/6) | 4 (3/1) | 1.81E-02 | SLC37A2 |
| *chr15:101879092-101881492* | 2401 | 9 | 10 (8/2) | 5 (2/3) | 3.42E-02 | PCSK6 |
| *chr15:74240329-74242377* | 2049 | 9 | 11 (5/6) | 4 (2/2) | 2.72E-02 | LOXL1 |
| *chr16:84361522-84377707* | 16186 | 9 | 12 (10/2) | 3 (1/2) | 1.96E-03 | WFDC1 |
| *chr3:89270562-89271140* | 579 | 9 | 10 (7/3) | 5 (5/0) | 4.35E-02 | EPHA3 |
| *chr17:62526889-62540745* | 13860 | 9 | 11 (6/5) | 4 (2/2) | 2.21E-02-4.95E-02 | CEP95, SMURF2 |
| *chr22:37575016-37578883* | 3869 | 8 | 9 (5/4) | 5 (5/0) | 3.07E-02-4.01E-02 | C1QTNF6 |
| *chr4:175231373-175250343* | 18974 | 8 | 10 (4/6) | 4 (3/1) | 4.28E-03-2.88E-02 | CEP44 |
| *chr16:69714340-69734578* | 20239 | 7 | 9 (6/3) | 4 (4/0) | 4.44E-02 | NFAT5 |
| *chr3:65605257-65608129* | 2873 | 7 | 8 (2/6) | 5 (4/1) | 4.57E-02 | MAGI1 |
| *chr3:89186869-89187058* | 190 | 7 | 8 (3/5) | 5 (5/0) | 2.38E-02 | EPHA3 |
| *chr3:184087831-184112540* | 24713 | 7 | 7 (5/2) | 6 (0/6) | 9.45E-03-3.58E-02 | THPO, CHRD |
| *chr6:36928408-36937172* | 8769 | 7 | 9 (6/3) | 4 (4/0) | 4.44E-02-4.44E-02 | PI16, MTCH1 |
| *chr11:14185641-14190574* | 4936 | 7 | 7 (2/5) | 5 (5/0) | 1.62E-02-4.89E-02 | SPON1 |
| *chr11:63491191-63518775* | 27586 | 7 | 9 (6/3) | 3 (3/0) | 2.63E-02-2.63E-02 | RTN3 |
| *chr19:19647199-19653445* | 6247 | 7 | 9 (5/4) | 3 (2/1) | 4.23E-02 | CILP2 |
| *chr21:45777972-45783129* | 5158 | 7 | 9 (6/3) | 3 (3/0) | 2.63E-02 | TRPM2 |

## Discussion

This is the first GWAS performed for cancer cachexia. True to the preliminary nature of the study, we identified CNV/CNVRs associated with cachexia at a nominal p-value cut-off (p-value <0.05) of marker associations. Hence, we have not applied the threshold for genome wide significance. This is consistent with the study design of previously published stage 1 studies utilizing GWAS approaches [17, 18]. Although candidate gene SNP studies identified certain loci associated with cachexia^10^, the unbiased genome-wide approach in the current study identified several new genes associated with weight loss in patients with cancer. Several genes embedded within common CNV/CNVRs were shown to be differentially expressed in skeletal muscle datasets available in the public domain, and were involved in various well-known pathways such as sphingolipid signaling, inflammatory pathways, Foxo signaling and Oncostatin M signaling^1,2,24^. Results from these CNV/CNVRs indicate that these germline variants show association with weight loss in patients with cancer and likely mediate the effects through gene dosage even though other potential non-coding regulatory mechanisms may also confer phenotypic changes. 83% of CNV/CNVRs were also present in external databases such as DGV and the 1000 Genomes project phase 3 data thereby adding credence to the findings that the profiled CNVs and their identified associations with weight loss are common polymorphisms in populations and may be explored for their potential value as biomarkers from germline DNA.

It is known that 10% of the human genome is composed of CNVs which may potentially alter gene dosage and therefore confer a phenotype^25^. Though further independent replication studies are warranted to confirm the CNV associations in a larger cohort of patients, the potential functional annotations of the embedded genes within CNVs/CNVRs in a human skeletal muscle tissue-specific context is unique to our study. When the CNV/CNVR embedded protein coding genes were subjected to pathway analysis, many known functions such as Oncostatin M signaling, JAK-STAT signaling, Foxo signaling were identified. Many of the functionally characterized cachexia genes such as JAK1, JAK2, LIFR, SMAD4, CAMK2B^26–28^ identified as CNVs in 5-15% frequency in this study were also reported in the reference populations (1000 Genomes Project and DGV), lending credence to the study findings despite the limited sample size. Of interest, exploring and targeting the JAK/STAT would be an interesting therapeutic option. Dysregulated JAK/STAT pathway in cancer causes muscle wasting through activation of STAT3^29^. Independent studies have shown that pharmacological intervention of JAK inhibitors reduced muscle wasting and leukemia inhibitory factor associated adipose loss in animal model of cachexia^30^. As JAK inhibitors are approved for myelofibrosis by FDA^31,32^, and ruxolitinib (JAK 1/2 inhibitor) being in phase 1 clinical trial for cachexia, it remains to be seen if these inhibitors can be used as a therapeutic option for cachexia. If JAK1, JAK2 and LIFR are validated in larger cachexia cohorts, it can potentially be considered as a genetic biomarker for cancer cachexia susceptibility.

Furthermore, to gain biological relevance for the identified CNVs, we mapped the CNV/CNVRs embedded genes (this study) to the previously reported differentially expressed genes from two independent human skeletal muscle gene expression datasets. The gene expression datasets utilized different profiling platforms (arrays and next generation sequencing) utilizing muscle biopsies from Oesophagogastric and pancreatic cancer patients. The CNVs/CNVRs identified in the current study utilized muscle biopsies from Oesophagogastric and lung cancer patients. Despite these differences, it is encouraging that the gene overlap across datasets associated with cancer associated muscle wasting is suggestive of potential functional role for germline CNV embedded genes expressed at the muscle tissue level.

As our understanding of cancer cachexia keeps evolving, applying the findings clinically in terms of identifying biomarkers remains a challenge. Considering the age and status of the patients who are diagnosed with cachexia, obtaining muscle biopsies is invasive when compared to collecting blood samples to identify biomarkers. The comparisons made in our study to assess skeletal muscle tissue gene expressions were from unmatched sample sets. Despite this limitation, several genes embedded within CNV/CNVRs could be interrogated due to the confidence in the CNV calls made across genotyping platforms in the DGV, 1000 Genomes Project and Affymetrix platform (current study) to identify common CNVs in diverse populations. Matched samples (blood and muscle biopsy from the same patient) and profiling for gene expressions and germline CNVs are needed to unequivocally identify expression Quantitative Traits Loci (CNV-eQTLs). Role of CNVs as genetic determinants of disease and/or trait susceptibility are now increasingly recognized in several diseases and traits^14,15,33^. Our studies potentially serve to illustrate that the CNV-GWAS as a premise in the domain of cancer cachexia is feasible.

Several circulating protein biomarkers have been identified as potential biomarkers for cachexia, which requires validation in independent cohorts^34,35^. It is possible that different primary cancers generate different circulating factors which makes the process of identifying a universal cachexia marker a challenge. One of the alternatives to identify cachexia biomarkers for early detection is the use of CNV as DNA biomarkers since germline DNA is known to remain stable across generations and not influenced by rearrangements and other chromosomal aberrations due to genomic instability at the level of somatic (cancer) genome.

Being proof of principle study, our sample size was limited. Understanding the genetic predisposition of cancer cachexia requires a collective effort to pool resources and can only be possible with collaborative efforts. This study may be seen as an example of a multicenter collaborative effort in collecting the samples along with clinical information. We used extremes of phenotypes by using weight stable cancer controls and cachectic cases with patients with cancer who had >10% weight loss. While the propensity of cachexia may differ among cancer types, it would be important to extend the study to different cancer types in future. The current study focus was CNVs (and their embedded coding genes), but SNP associations with the cachexia phenotype at a whole genome level should be explored in future studies using larger sample sizes in a multistage design to identify and validate SNP markers. SNP markers in the array we used are largely in the intergenic regions and fine mapping is warranted to identify putative causal loci.

In conclusion, this is the first collaborative effort at an international level to perform genome-wide association studies to identify genetic variants for cancer cachexia. Validation of these results in independent cohorts is warranted.

## Data Availability

All data produced in the present work are contained in the manuscript

## Acknowledgements

This article is dedicated to Professor. Kenneth Fearon who encouraged cancer cachexia genetic predisposition studies.

This work was funded through operating grants from Canadian Institutes of Health Research (CIHR) to SD and VB and through a grant from the Terry Fox Research Institute (TFRI), Canada (BG)

The authors comply with the ethical guidelines for authorship and publishing in the Journal of Cachexia, Sarcopenia and Muscle^36^.

## Supporting information

**Supplementary Table S1**. Complete list of significant CNV/CNVRs: 896 CNV/CNVRs with p<0.05 along with their frequency is represented. Total aberrations indicate the total number of aberrations in cases and controls combined. Gain frequency in cases and controls were calculated using the the total number of aberrations in cases (n=71) and controls (n=103), respectively. The presence of CNV/CNVRs from this study were mapped to curated databases such as 1000 genomes and database of genomic variants. CNVs/CNVRs highlighted in grey are presented in Table 2 as CNVs/CNVRs showing the highest frequencies in the study cohort with roles in cancer and not in cancer cahexia. Genes which are not annotated or for which orthologs are not yet identified were not taken into consideration for representation of the highest frequencies shown in grey.

**Supplementary Table S2**. Complete list of genes embedded within CNVs/CNVRs shown to be differentially expressed in skeletal muscle of patients with cancer cachexia versus no cachexia.

